# Telomere length and its associations with mental disorders, age and genetic risk for mental disorders

**DOI:** 10.1101/2022.03.29.22273140

**Authors:** Julian Mutz, Cathryn M Lewis

## Abstract

**Background:** Mental disorders are associated with substantially increased morbidity and reduced life expectancy. Accelerated biological ageing might contribute to excess mortality of individuals with mental disorders. The aim of this study was to characterise telomere length, a biological hallmark of ageing, in individuals with mental disorders, and to examine associations between telomere length, age and genetic risk for mental disorders.

**Methods:** The UK Biobank is a multicentre, community-based observational study that recruited >500,000 middle-aged and older adults across England, Scotland and Wales. Average leukocyte telomere length (T/S ratio) was measured using quantitative polymerase chain reaction. Polygenic risk scores (PRS) were calculated for individuals of European ancestry. We estimated differences in T/S ratio and age-related changes in T/S ratio between individuals with anxiety disorder, depression or bipolar disorder and people without mental disorders. We also estimated associations between T/S ratio and PRS for these three disorders.

**Results:** The analyses included up to 308,725 participants. Individuals with depression had shorter telomeres than people without mental disorders (adjusted *β* = -0.011, 95% CI -0.019 to -0.004, *p*_Bonf._ = 0.027). There was only limited evidence of case-control differences in telomere length for anxiety disorders or bipolar disorders. Age-related changes in telomere length did not differ between individuals with and without mental disorders. PRS for depression were associated with shorter telomeres (*β* = -0.006, 95% CI -0.010 to -0.003, *p*_Bonf._ = 0.001). There was no evidence that PRS for anxiety disorder or bipolar disorder were associated with telomere length.

**Conclusion:** Although telomere length is a biological hallmark of ageing, we observed limited evidence that leukocyte telomere length is a clinically useful marker to quantify accelerated biological ageing in middle-aged and older adults with a lifetime history of anxiety disorder, depression or bipolar disorder.

## Introduction

Telomeres are repetitive nucleoprotein complexes at the chromosome ends that play an important role in maintaining genomic stability. Telomeres shorten with each cell division and therefore represent a biological marker of replicative history and cellular age.^1,2^ Although telomere length is highly heritable,^3^ age-related attrition results from biological and environmental factors, including lifestyle and chronic stress.^4^ Telomere attrition has been associated with an increased risk of age-related diseases. Mendelian randomisation analyses in the UK Biobank, a major biomedical database, suggested that telomere length had widespread influence on biomedical traits, disease risk, multiple body systems and life expectancy.^5^

Individuals with mental disorders have an increased prevalence of age-related diseases and a lower life expectancy.^6^ They also show signs of accelerated biological ageing, including advanced brain ageing,^7^ changes in DNA methylation,^8^ greater levels of inflammation,^9^ elevated frailty^10^ and differences in physiological measures such as grip strength.^11-13^ Telomere length as a molecular marker of biological senescence could provide insight into the relationship between mental health and accelerated biological ageing. Data from a meta-analysis suggested that individuals with anxiety disorders, depressive disorders and posttraumatic stress disorder had shorter telomeres than people without these disorders.^14^ Findings regarding bipolar disorder have been inconsistent,^15^ with some studies observing longer telomeres in patients,^14^ likely due to lithium treatment.^16^ Bipolar disorder patients not exposed to lithium had shorter telomeres than patients who had been treated with lithium.^17^

Most previous studies have had limited sample sizes and few studies have included cross-disorder comparisons within the same database. There has also been little exploration of associations between telomere length and genetic risk for mental disorders. The UK Biobank provides an unprecedented resource to investigate health and ageing trajectories, with the world’s largest database of leukocyte telomere length measurements. The aim of this study was to characterise telomere length in individuals with a history of anxiety disorder, depression or bipolar disorder, and to examine associations between telomere length, age and polygenic risk scores for these disorders.

## Methods

### Study population

The UK Biobank is a prospective study of >500,000 UK residents aged 37–73 at baseline who were recruited between 2006 and 2010. The study rationale and design have been described elsewhere.^18^ Briefly, individuals registered with the UK National Health Service (NHS) and living within a 25-mile (∼40 km) radius of one of 22 assessment centres were invited to participate. Participants provided data on their sociodemographic characteristics, health behaviours and medical history, underwent physical examination and had blood and urine samples taken. Linked hospital inpatient records are available for most participants and primary care data are available for half of the participants. A third of participants completed an online follow-up mental health questionnaire (MHQ) between 2016 and 2017.

### Leukocyte telomere length

Details of the measurement of leukocyte telomere length (UK Biobank data fields 22191 and 22192), including technical adjustments, have been reported elsewhere.^19^ Briefly, relative telomere length was measured using a validated quantitative polymerase chain reaction (PCR) assay that expresses telomere length as the ratio of the telomere repeat copy number relative to the reference single-copy hemoglobin subunit beta gene (T/S ratio). This ratio is proportional to the individual’s average telomere length.^20^ Measurements were adjusted for operational and technical parameters (PCR machine, staff member, enzyme batch, primer batch, temperature, humidity, primer batch × PCR machine, primer batch × staff member, A260/A280 ratio of the DNA sample, and A260/A280 ratio squared), log_e_ transformed and Z-standardised. T/S ratio was also converted to base pairs using the formula: base pairs = 3274+2413×((T/S-0.0545)/1.16).^21^

### Mental disorders

We identified individuals with lifetime anxiety disorders, depression or bipolar disorder using our previously reported criteria.^11-13^ Data sources included the modified Composite International Diagnostic Interview Short Form (CIDI-SF), self-report questions on (hypo)mania and a question on psychiatric diagnoses (field 20544) which were assessed as part of the MHQ, the nurse-led baseline interview in which participants reported medical diagnoses (field 20002), hospital inpatient records (ICD-10 codes), primary care records (Read v2 or CTV3 codes) and self-report questions on mood disorders from the baseline assessment (field 20126). Individuals with psychosis were excluded from all groups and individuals with bipolar disorder were excluded from the anxiety disorder group due to their increased risk of physical multimorbidity.^22,23^ The depression and bipolar disorder groups were mutually exclusive. Individuals could be included in both the anxiety disorder and the depression group.

Individuals in the control group had no mental disorders: (i) did not report “schizophrenia”, “depression”, “mania / bipolar disorder / manic depression”, “anxiety / panic attacks”, “obsessive compulsive disorder”, “anorexia / bulimia / other eating disorder”, “post-traumatic stress disorder” at the baseline interview; (ii) no psychiatric diagnoses on the MHQ; (iii) no current psychotropic medication use at baseline (field 20003);^24^ (iv) no ICD-10 Chapter V code in their hospital inpatient record (F20-F99), except for organic causes or substance use; (v) no primary care record containing diagnostic codes for mental disorders;^25^ (vi) not classified as individuals with probable mood disorder; (vii) no Patient Health Questionnaire-9 (PHQ-9) or Generalised Anxiety Disorder Assessment (GAD-7) sum score of ≥5; (viii) never felt worried, tense, or anxious for most of a month or longer (field 20421); (ix) not identified as cases based on the CIDI-SF and questions on (hypo)manic symptoms.^12,13^

### Genetic quality control

Genetic quality control was performed as described previously.^26^ Individuals were excluded where recommended by the UK Biobank for unusual levels of missingness (>5%) or heterozygosity.^18^ Using the genotyped single nucleotide polymorphisms (SNPs), individuals with call rate of less than 98%, who were genetically related to another individual in the dataset (KING *r* < 0.044, equivalent to removing third-degree relatives and closer)^27^ or whose self-reported and genotypic sex did not match (X-chromosome homozygosity (*F*_X_) < 0.9 for phenotypic males, *F*_X_ > 0.5 for phenotypic females) were also excluded. To account for familial correlation, removal of relatives was performed using a “greedy” algorithm, which minimises exclusions (for example, by excluding the child in a mother–father–child trio). All analyses were limited to individuals of European ancestry, as defined by 4-means clustering on the first two genetic principal components (PCs) provided by the UK Biobank.^28^ Principal components analysis was also performed on the European-only subset of the data using FlashPCA2.^29^

### Polygenic risk scores

Polygenic risk scores (PRS) for anxiety disorder, depression and bipolar disorder were calculated using PRSice v.2.^30^ This method involves calculating PRS as the sum of risk alleles weighted by SNP effect sizes from independent genome-wide association study (GWAS) summary statistics (Supplementary Table 1). Clumping was performed to remove SNPs in high linkage disequilibrium (defined as *r*^2^ ≥ 0.1 within 250 kilobases on each side) as linkage disequilibrium can falsely inflate polygenic scores. PRS were calculated at 11 *p*-value thresholds (5×10^−8^, 1×10^−5^, 1×10^−3^, 0.01, 0.05, 0.1, 0.2, 0.3, 0.4, 0.5 and 1) and the PRS most predictive threshold was selected for regression analyses. All individual-level PRS were standardized prior to analyses.

### Covariates

Covariates were identified from previous research and included age, sex, white blood cell count, Townsend deprivation index, physical activity, smoking status, body mass index, body fat percentage and C-reactive protein. For the analyses of PRS, covariates included the first six ancestry-informative population PCs, batch number and assessment centre.

### Statistical analyses

Regression analyses were performed in R (version 3.6.2).

Sample characteristics were summarised using means and standard deviations or counts and percentages. Differences in T/S ratio (log z adjusted) between individuals with anxiety disorder, depression or bipolar disorder and the control group were estimated using ordinary least squares regression (± 95% confidence intervals). For these analyses, we fitted minimally adjusted models that included age and sex and fully adjusted models that included all covariates. Finally, associations between T/S ratio (log z adjusted) and PRS for anxiety, depression and bipolar disorder were estimated using ordinary least squares regression. These models included six PCs, batch number and assessment centre.

We calculated adjusted *P*-values to correct for multiple testing. Two methods were used: (1) Bonferroni and (2) Benjamini & Hochberg,^31^ all two-tailed, with *α* = .05 and a false discovery rate of 5%, respectively. *P*-values were corrected for six to eight tests (see tables for details). We have opted for this approach because the Bonferroni correction may be too conservative and potentially leads to a high number of false negatives.

### Additional analyses

We repeated our main analyses (i) with the bipolar disorder group stratified by current lithium use and (ii) to assess the association between PRS independent of diagnosis and treatment-related confounders, we stratified the PRS analyses by case status. As a sensitivity analysis, we excluded individuals with comorbid depression and anxiety disorder. Finally, we repeated the main and sensitivity analyses by antidepressant medication use.

## Results

After quality control exclusions and restricting our sample to individuals of European ancestry, 458,078 participants (out of 502,476) had data on both telomere length and polygenic risk scores. We retained up to 308,725 participants with complete data on all covariates. 41,524 individuals had lifetime anxiety disorders, 84,965 had lifetime depression and 3449 had bipolar disorders (Table 1).

**Table 1.** Sample characteristics of individuals with mental disorders and non-psychiatric controls

|  | **Anxiety disorder**<br>N=41524 | **Depression**<br>N=84965 | **Bipolar disorder**<br>N=3449 | **Controls**<br>N=223760 |
| --- | --- | --- | --- | --- |
| **T/S ratio** (log z adjusted) |  |  |  |  |
| Mean (SD) | 0.02 (0.99) | 0.02 (0.99) | 0.01 (1.01) | -0.01 (1.00) |
| **Telomere length** (base pairs) |  |  |  |  |
| Mean (SD) | 4897.74 (271.66) | 4896.79 (270.31) | 4895.22 (270.45) | 4888.25 (271.99) |
| **Age** |  |  |  |  |
| Mean (SD) | 56.04 (7.86) | 55.55 (7.88) | 54.76 (7.97) | 56.77 (8.08) |
| **Sex** |  |  |  |  |
| Female | 27316 (65.8%) | 55322 (65.1%) | 1915 (55.5%) | 111064 (49.6%) |
| Male | 14208 (34.2%) | 29643 (34.9%) | 1534 (44.5%) | 112696 (50.4%) |
| **Neighbourhood deprivation** |  |  |  |  |
| Mean (SD) | -1.36 (3.04) | -1.22 (3.06) | -0.71 (3.21) | -1.68 (2.87) |
| **Walking<sup>1</sup>** |  |  |  |  |
| Mean (SD) | 5.31 (1.98) | 5.32 (1.99) | 5.37 (2.05) | 5.43 (1.91) |
| **Moderate activity<sup>1</sup>** |  |  |  |  |
| Mean (SD) | 3.53 (2.35) | 3.52 (2.36) | 3.64 (2.42) | 3.66 (2.31) |
| **Vigorous activity<sup>1</sup>** |  |  |  |  |
| Mean (SD) | 1.74 (1.91) | 1.74 (1.92) | 1.87 (2.03) | 1.92 (1.96) |
| **Smoking status** |  |  |  |  |
| Never | 21393 (51.5%) | 42648 (50.2%) | 1548 (44.9%) | 125978 (56.3%) |
| Former | 15494 (37.3%) | 31614 (37.2%) | 1248 (36.2%) | 77482 (34.6%) |
| Current | 4637 (11.2%) | 10703 (12.6%) | 653 (18.9%) | 20300 (9.1%) |
| **Body mass index** |  |  |  |  |
| Mean (SD) | 27.27 (4.99) | 27.63 (5.11) | 28.02 (5.33) | 27.14 (4.45) |
| **Body fat percentage** |  |  |  |  |
| Mean (SD) | 32.68 (8.54) | 32.94 (8.61) | 31.90 (8.80) | 30.45 (8.34) |
| **Obesity** |  |  |  |  |
| Underweight, BMI < 18.5 | 281 (0.7%) | 442 (0.5%) | 17 (0.5%) | 963 (0.4%) |
| Normal, 18.5 ≤ BMI < 25 | 14593 (35.1%) | 27774 (32.7%) | 1052 (30.5%) | 75326 (33.7%) |
| Overweight, 25 ≤ BMI < 30 | 16684 (40.2%) | 34299 (40.4%) | 1346 (39.0%) | 98576 (44.1%) |
| Obese, 30 ≤ BMI < 35 | 6959 (16.8%) | 15335 (18.0%) | 709 (20.6%) | 36773 (16.4%) |
| Severely obese, BMI ≥ 35 | 3007 (7.2%) | 7115 (8.4%) | 325 (9.4%) | 12122 (5.4%) |
| **White blood cell count** |  |  |  |  |
| Mean (SD) | 6.90 (1.92) | 6.96 (1.96) | 7.18 (2.47) | 6.81 (2.03) |
| **C-reactive protein** |  |  |  |  |
| Mean (SD) | 2.60 (4.26) | 2.69 (4.31) | 2.80 (4.12) | 2.41 (4.14) |
| **Antidepressant use** |  |  |  |  |
| No | 32645 (78.6%) | 66508 (78.3%) | 2408 (69.8%) | 223760 (100.0%) |
| Yes | 8879 (21.4%) | 18457 (21.7%) | 1041 (30.2%) | 0 (0.0%) |
| **Antipsychotic use** |  |  |  |  |
| No | 41293 (99.4%) | 84592 (99.6%) | 3204 (92.9%) | 223760 (100.0%) |
| Yes | 231 (0.6%) | 373 (0.4%) | 245 (7.1%) | 0 (0.0%) |
| **Lithium use** |  |  |  |  |
| No | 41483 (99.9%) | 84849 (99.9%) | 3144 (91.2%) | 223760 (100.0%) |
| Yes | 41 (0.1%) | 116 (0.1%) | 305 (8.8%) | 0 (0.0%) |

Note: SD = standard deviation; BMI = body mass index. Units: white blood cell count, $\times 10^9$ cells/litre; c-reactive protein, milligrams/litre; body mass index, kilograms/metres<sup>2</sup>. <sup>1</sup> number of days per week engaging in these activities for 10+ minutes continuously.

Average telomere length (T/S ratio log z adjusted) was slightly greater in individuals with mental disorders relative to the non-psychiatric control group (Supplement Figure 1). However, this difference could be explained by group differences in age and sex. After adjusting for age and sex (Model 1) and other potential confounders (Model 2), we observed that individuals with mental disorders had slightly shorter telomeres (Figure 1). This difference was only statistically significant for the comparison between individuals with depression and the control group (fully adjusted *β* = -0.011, 95% CI -0.019 to -0.004, *p*_Bonf._ = 0.027) (Table 2).

**Table 2.** T/S ratio (log z adjusted) in individuals with mental disorders.

| Term | Model 1 |  |  |  |  | Model 2 |  |  |  |  |
| --- | --- | --- | --- | --- | --- | --- | --- | --- | --- | --- |
| | $\beta$ | 95% CI | | $p_{\text{Bonf.}}$ | $p_{\text{BH}}$ | $\beta$ | 95% CI | | $p_{\text{Bonf.}}$ | $p_{\text{BH}}$ |
| Controls | Ref | - | - | - | - | Ref | - | - | - | - |
| Anxiety disorder | -0.010 | -0.020 | 0.001 | 0.396 | 0.099 | -0.003 | -0.013 | 0.008 | >0.999 | 0.609 |
| Depression | -0.023 | -0.030 | -0.015 | <0.001 | <0.001 | -0.011 | -0.019 | -0.004 | 0.027 | 0.014 |
| Bipolar disorder | -0.033 | -0.065 | 0.000 | 0.303 | 0.099 | -0.011 | -0.044 | 0.022 | >0.999 | 0.601 |
| No Lithium | -0.045 | -0.079 | -0.011 | 0.082 | 0.027 | -0.024 | -0.058 | 0.010 | >0.999 | 0.193 |
| Lithium | 0.093 | -0.016 | 0.203 | 0.754 | 0.126 | 0.121 | 0.011 | 0.230 | 0.243 | 0.061 |

*Note:* $\beta$ = ordinary least squares regression beta coefficient; CI = confidence interval; Ref = reference group; Bonf. = Bonferroni; BH = Benjamini & Hochberg. Model 1 – adjusted for age and sex; Model 2 – adjusted for age, sex, white blood cell count, Townsend deprivation index, physical activity, smoking status, body mass index, body fat percentage and C-reactive protein. *P*-values corrected for six (main analysis) and eight (bipolar disorder cases stratified by lithium use) tests.

**Figure 1.**
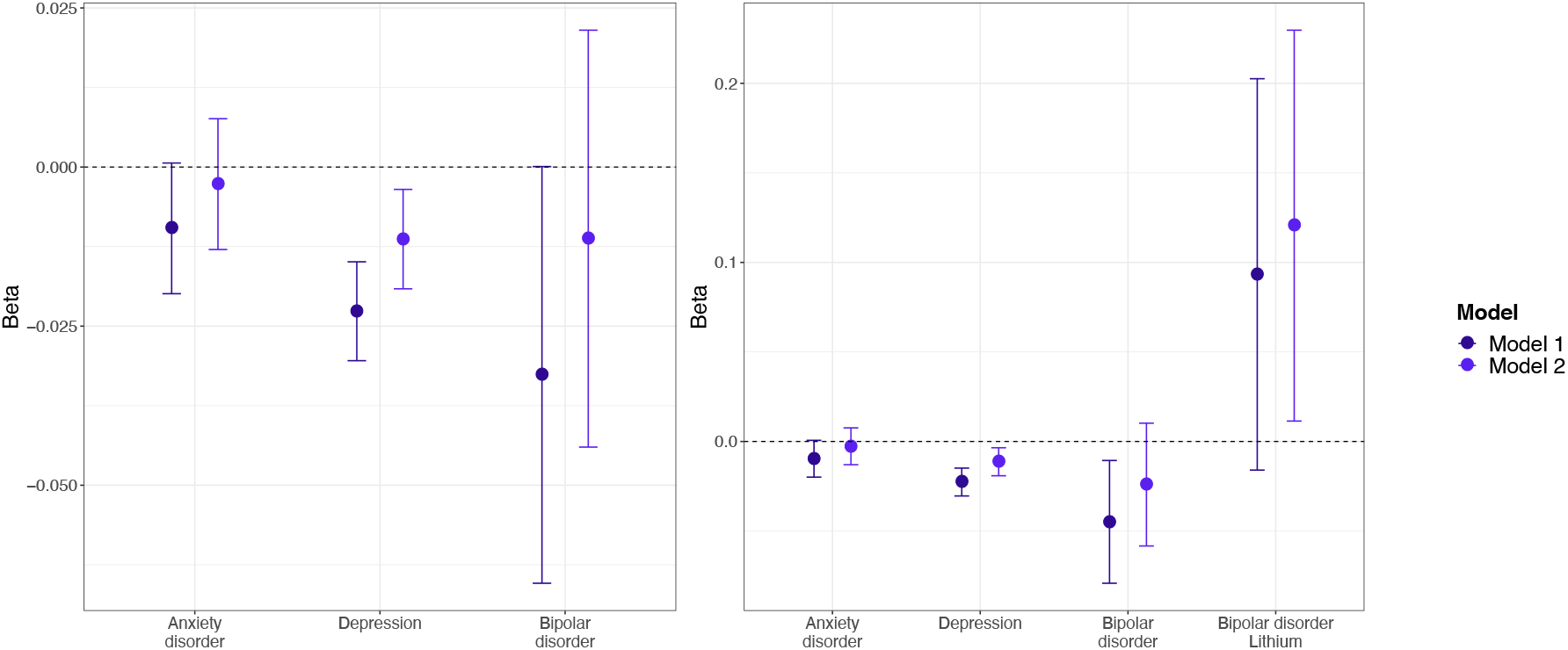
Average T/S ratio (log z adjusted) in individuals with mental disorders compared to non-psychiatric controls (reference group). Estimates shown are ordinary least squares regression beta coefficients and 95% confidence intervals. Model 1 – adjusted for age and sex; Model 2 – adjusted for age, sex, white blood cell count, Townsend deprivation index, physical activity, smoking status, body mass index, body fat percentage and C-reactive protein.

When stratifying individuals with bipolar disorder by current lithium use, we found that after adjusting for age and sex, telomeres were shorter in individuals who did not use lithium (adjusted *β* = -0.045, 95% CI -0.079 to -0.011, *p*_BH_ = 0.027), relative to the control group. Individuals with bipolar disorder who used lithium had slightly longer telomeres than individuals in the control group (fully adjusted *β* = 0.121, 95% CI 0.011 to 0.230, *p*_BH_ = 0.061), although this difference was not statistically significant after multiple testing correction. Individuals with anxiety disorder or depression who reported antidepressant medication use had shorter telomeres than the control group (Supplement Figure 2 and Supplement Table 2).

**Figure 2.**
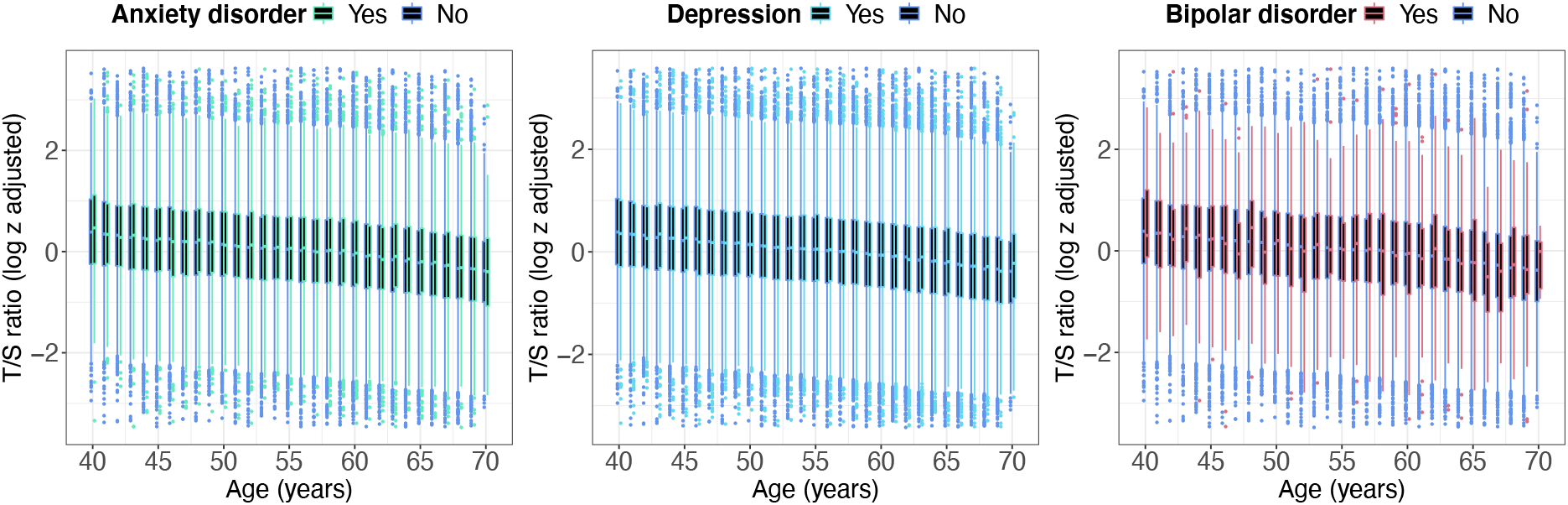
Age-related changes in average T/S ratio (log z adjusted) in individuals with mental disorders and non-psychiatric controls. T/S ratio values below the 0.01st or above the 99.99th percentile not shown.

As expected, telomere length reduced with age (Figure 2). There was no evidence that age-related changes in T/S ratio differed between individuals with mental disorders and people without mental disorders.

The distribution of polygenic risk scores in individuals with mental disorders and the non-psychiatric control group are shown in Supplement Figure 3, confirming that small to moderate increased PRS in individuals with mental disorders. There was little evidence of an association between the PRS for anxiety disorder, depression or bipolar disorder and telomere length (Supplement Figure 4). In a regression model, the PRS for depression was associated with shorter telomeres (adjusted *β* = -0.006, 95% CI -0.010 to -0.003, *p*_Bonf._ = 0.001). However, the magnitude of this association was negligible. There was no evidence that the PRS for anxiety disorder or bipolar disorder were associated with telomere length (Figure 3). When stratifying these analyses by case status, the PRS for depression was only associated with shorter telomeres in individuals without mental disorders (Supplement Table 3).

**Figure 3.**
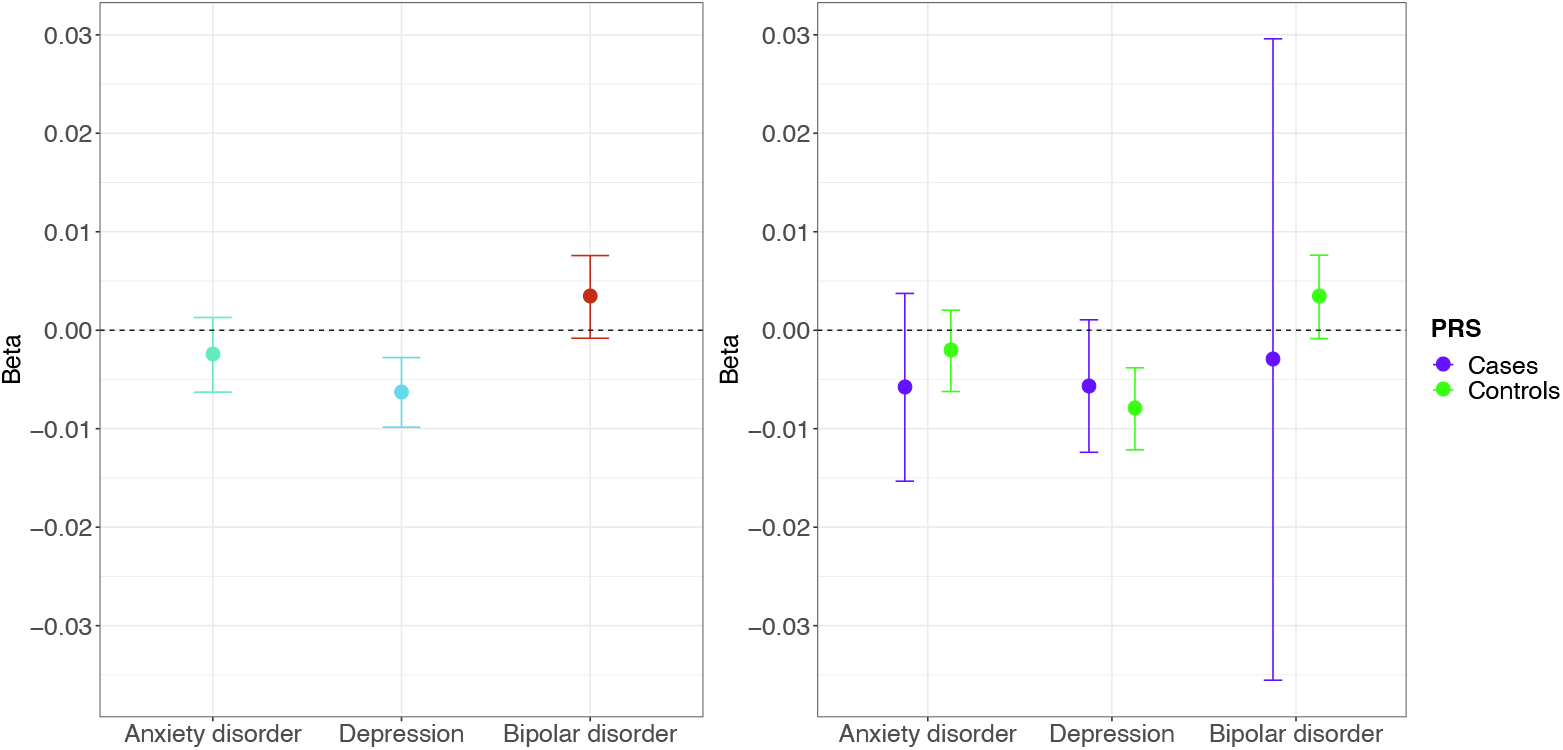
Associations between average T/S ratio (log z adjusted) and polygenic risk scores for anxiety disorder, depression and bipolar disorder. All analyses were adjusted for the first six ancestry-informative population principal components, batch number and assessment centre.

### Sensitivity analysis

53,780 individuals had a history of depression without comorbid anxiety disorders, while 14,829 individuals had anxiety disorders without comorbid depression. Individuals with depression had shorter telomeres (fully adjusted *β* = -0.015, 95% CI -0.025 to -0.006, *p*_Bonf._ = 0.004) (Supplement Figure 5), and this difference was slightly greater than in the main analysis. There was no evidence of a difference in telomere length between individuals with anxiety disorders and the non-psychiatric control group (fully adjusted *β* = 0.004, 95% CI -0.012 to 0.021, *p*_Bonf._ > 0.999) (Supplement Table 4).

Finally, individuals with depression who reported antidepressant medication use had shorter telomeres than individuals with depression not reporting medication use, relative to the control group (Supplement Figure 6 and Supplement Table 5). There was no evidence of an association between telomere length and anxiety disorder status, irrespective of antidepressant medication use.

## Discussion

Individuals with depression had slightly shorter telomeres than people without mental disorders. There was only limited evidence that telomere length differed between individuals with anxiety disorder or bipolar disorder and people without mental disorders. Notably, lithium use was associated with elongated telomeres in individuals with bipolar disorder. Antidepressant medication use was associated with reduced telomere length in individuals with depression. Age-related changes in telomere length did not differ between individuals with and without mental disorders. Polygenic risk scores for depression were associated with shorter telomeres. There was no evidence that polygenic risk scores for anxiety disorder or bipolar disorder were associated with telomere length.

The observation that depression was associated with shorter telomeres in the UK Biobank is consistent with data from meta-analyses.^14,32,33^ Although meta-analyses also provided evidence of an association between anxiety disorders and shorter telomeres,^14,34^ we did not observe a statistically significant difference between individuals with anxiety disorders and people without mental disorders. This discrepancy could be due to adjustment for multiple potential confounders in the present study. Although many previous studies examining telomere length in individuals with mental disorders adjusted for age and sex, most did not adjust for other confounders. Data from two meta-analyses found no association between bipolar disorder and telomere length.^14,35^ However, the most recent meta-analysis suggested that bipolar disorder patients had shorter telomeres compared to participants in the control group.^36^ Inconsistencies between studies could relate to differences in sample characteristics. For example, a recent study found that patients with bipolar disorder type I, but not bipolar disorder type II, had shorter telomeres than healthy controls.^37^ Another study did not observe group differences in telomere length between bipolar disorder subtypes but was likely underpowered (*n*=119 vs *n*=12, respectively).^17^ The possibility of subgroup differences should be further investigated.

Our finding that lithium use modified the direction of association between bipolar disorder and telomere length is consistent with previous observations that lithium treatment was associated with increased telomere length^38,39^ and that telomere length positively correlated with duration of lithium treatment^16,38^. A recent study found that bipolar disorder patients who had never been treated with lithium had shorter telomeres than healthy controls, while patients treated with lithium had longer telomeres than the never treated patients, although not compared to healthy controls.^17^ Our finding that antidepressant medication use was associated with reduced telomere length in individuals with depression confirms a preliminary study showing that antidepressant use (*n*=40) was associated with shorter telomeres, independent of depression diagnosis and current depression severity.^40^

Although previous research suggested that age-related decline in telomere length was greater in individuals with chronic stress or comorbidities,^41^ we observed similar association between telomere length and age in individuals with mental disorders and the control group.

Multiple studies have examined polygenic scores for telomere length to predict mental disorders,^42^ but there has been limited research on polygenic risk scores for mental disorders to predict telomere length. Preliminary studies found that unaffected first-degree relatives of individuals with bipolar disorder had shorter telomeres than healthy controls.^39,43^ Similarly, a small cross-sectional study found that daughters of mothers with depression had shorter telomeres than daughters of never depressed mothers.^44^ Although these findings suggest that increased genetic risk for mental disorders may affect telomere length, these studies were limited by modest sample sizes and cannot fully disentangle genetic and environmental risk factors. Depression polygenic risk scores were not associated with telomere length or telomere attrition rate in 2032 adults aged 18 to 65 years.^42^ A study of 290 adults without depression also found no evidence that polygenic risk scores for depression, bipolar disorder or schizophrenia were associated with telomere length.^40^ We found that polygenic risk scores for depression, but not for anxiety disorder or bipolar disorder, were associated with shorter telomeres, although the strength of this association was negligible. Our finding that depression polygenic risk scores were associated with telomere length only in individuals without mental disorders could be explained by the lower sample size in the case group and warrants replication.

In contrast to most previous studies, we examined associations between telomere length and mental disorders in the same data using a shared control group, allowing for cross-disorder comparison. Our study also included a considerably larger number of individuals with mental disorders. Indeed, the UK Biobank is by far the largest data resource with measured telomere length, which allowed us to adjust for a range of potential confounders.

Our observational study has limitations. The age range was limited to middle-aged and older adults (most between 40 to 69 years old). A previous Dutch study found no difference in telomere length between individuals with current depression aged 60 or older compared to never depressed individuals, suggesting that our findings might not extrapolate to late-life depression.^45^ For the cross-sectional analyses, our ability to draw causal conclusions was limited. Longitudinal studies have found that major depressive disorder^46^ or persistent internalizing disorders in men^47^ predicted reduced telomere length, although not all studies found evidence of a prospective association between mental disorders and telomere length.^33,48^ A large study of 67,306 individuals aged 20-100 from the Danish general population found no evidence that telomere length predicted depression prospectively or that genetically shorter telomeres predicted depression. Nevertheless, the authors observed that depression was associated with shorter telomeres cross-sectionally, which could be explained by depression causing shorter telomeres or residual confounding.^49^ Finally, telomeres were measured from leukocyte DNA and findings might differ when examining other tissues. However, research suggests that leukocyte telomeres correlate well with telomere length measured in other tissues.^50^

## Conclusion

Although telomere length is a biological hallmark of ageing, we observed limited evidence that leukocyte telomere length is a clinically useful marker to quantify accelerated biological ageing in middle-aged and older adults with a lifetime history of anxiety disorder, depression or bipolar disorder in the UK Biobank.

## Data Availability

The data used are available to all bona fide researchers for health-related research that is in the public interest, subject to an application process and approval criteria. Study materials are publicly available online at http://www.ukbiobank.ac.uk.

## Ethics

Ethical approval for the UK Biobank study has been granted by the National Information Governance Board for Health and Social Care and the NHS North West Multicentre Research Ethics Committee (11/NW/0382). No project-specific ethical approval is needed.

## Authorship contributions

JM conceived the idea of the study, acquired the data, carried out the statistical analysis, interpreted the findings, wrote the manuscript and revised the manuscript for final submission. CML acquired the studentship funding, interpreted the findings and critically reviewed the manuscript. Both authors read and approved the final manuscript.

## Funding and disclosure

JM receives studentship funding from the Biotechnology and Biological Sciences Research Council (BBSRC) (ref: 2050702) and Eli Lilly and Company Limited. CML is part-funded by the National Institute for Health Research (NIHR) Maudsley Biomedical Research Centre at South London and Maudsley NHS Foundation Trust and King’s College London. The views expressed are those of the authors and not necessarily those of the NHS, the NIHR or the Department of Health and Social Care. CML is a member of the Scientific Advisory Board of Myriad Neuroscience.

## Acknowledgments

This research has been conducted using data from UK Biobank, a major biomedical database. This project made use of time on Rosalind HPC, funded by Guy’s & St Thomas’ Hospital NHS Trust Biomedical Research Centre (GSTT-BRC), South London & Maudsley NHS Trust Biomedical Research Centre (SLAM-BRC), and Faculty of Natural Mathematics & Science (NMS) at King’s College London. Data access permission has been granted under UK Biobank application 45514.

## Supplementary material

Supplementary information is available online.

## Notes

### Competing Interest Statement

JM receives studentship funding from the Biotechnology and Biological Sciences Research Council (BBSRC) and Eli Lilly and Company Limited. CML is a member of the Scientific Advisory Board of Myriad Neuroscience.

## References

1 Harley, C. B., Futcher, A. B. & Greider, C. W. Telomeres shorten during ageing of human fibroblasts. Nature 345, 458–460, doi:10.1038/345458a0 (1990).

2 Allsopp, R. C. et al. Telomere length predicts replicative capacity of human fibroblasts. Proceedings of the National Academy of Sciences 89, 10114–10118, doi:10.1073/pnas.89.21.10114 (1992).

3 Broer, L. et al. Meta-analysis of telomere length in 19 713 subjects reveals high heritability, stronger maternal inheritance and a paternal age effect. European Journal of Human Genetics 21, 1163–1168, doi:10.1038/ejhg.2012.303 (2013).

4 Puterman, E., Lin, J., Krauss, J., Blackburn, E. H. & Epel, E. S. Determinants of telomere attrition over 1 year in healthy older women: stress and health behaviors matter. Molecular Psychiatry 20, 529–535, doi:10.1038/mp.2014.70 (2015).

5 Codd, V. et al. Polygenic basis and biomedical consequences of telomere length variation. medRxiv (2021).

6 Plana-Ripoll, O. et al. A comprehensive analysis of mortality-related health metrics associated with mental disorders: a nationwide, register-based cohort study. The Lancet 394, 1827–1835, doi:https://doi.org/10.1016/S0140-6736(19)32316-5 (2019).

7 Han, L. K. M. et al. Brain aging in major depressive disorder: results from the ENIGMA major depressive disorder working group. Molecular Psychiatry 26, 5124–5139, doi:10.1038/s41380-020-0754-0 (2021).

8 Jansen, R. et al. An integrative study of five biological clocks in somatic and mental health. eLife 10, e59479, doi:10.7554/eLife.59479 (2021).

9 Pitharouli, M. C. et al. Elevated C-Reactive Protein in Patients With Depression, Independent of Genetic, Health, and Psychosocial Factors: Results From the UK Biobank. American Journal of Psychiatry 178, 522–529, doi:10.1176/appi.ajp.2020.20060947 (2021).

10 Mutz, J., Choudhury, U., Zhao, J. & Dregan, A. Frailty in individuals with depression, bipolar disorder and anxiety disorders: longitudinal analyses of all-cause mortality. medRxiv, 2022.2002.2023.22271065, doi:10.1101/2022.02.23.22271065 (2022).

11 Mutz, J., Hoppen, T. H., Fabbri, C. & Lewis, C. M. Anxiety disorders and age-related changes in physiology. The British Journal of Psychiatry, 1–10, doi:10.1192/bjp.2021.189 (2022).

12 Mutz, J. & Lewis, C. M. Lifetime depression and age-related changes in body composition, cardiovascular function, grip strength and lung function: sex-specific analyses in the UK Biobank. Aging 13, 17038–17079 (2021).

13 Mutz, J., Young, A. H. & Lewis, C. M. Age-related changes in physiology in individuals with bipolar disorder. Journal of Affective Disorders 296, 157–168, doi:https://doi.org/10.1016/j.jad.2021.09.027 (2022).

14 Darrow, S. M. et al. The Association Between Psychiatric Disorders and Telomere Length: A Meta-Analysis Involving 14,827 Persons. Psychosomatic Medicine 78, 776–787, doi:10.1097/psy.0000000000000356 (2016).

15 Fries, G. R. et al. Accelerated aging in bipolar disorder: A comprehensive review of molecular findings and their clinical implications. Neuroscience & Biobehavioral Reviews 112, 107–116, doi:https://doi.org/10.1016/j.neubiorev.2020.01.035 (2020).

16 Coutts, F. et al. The polygenic nature of telomere length and the anti-ageing properties of lithium. Neuropsychopharmacology 44, 757–765, doi:10.1038/s41386-018-0289-0 (2019).

17 Pisanu, C. et al. Differences in telomere length between patients with bipolar disorder and controls are influenced by lithium treatment. Pharmacogenomics 21, 533–540, doi:10.2217/pgs-2020-0028 (2020).

18 Bycroft, C. et al. The UK Biobank resource with deep phenotyping and genomic data. Nature 562, 203–209 (2018).

19 Codd, V. et al. A major population resource of 474,074 participants in UK Biobank to investigate determinants and biomedical consequences of leukocyte telomere length. medRxiv (2021).

20 Lai, T.-P., Wright, W. E. & Shay, J. W. Comparison of telomere length measurement methods. Philosophical Transactions of the Royal Society B: Biological Sciences 373, 20160451 (2018).

21 Verhoeven, J. E. et al. Major depressive disorder and accelerated cellular aging: results from a large psychiatric cohort study. Molecular Psychiatry 19, 895–901, doi:10.1038/mp.2013.151 (2014).

22 Stubbs, B. et al. Physical multimorbidity and psychosis: comprehensive cross sectional analysis including 242,952 people across 48 low-and middle-income countries. BMC Medicine 14, 1–12 (2016).

23 Smith, D. J. et al. Multimorbidity in bipolar disorder and undertreatment of cardiovascular disease: a cross sectional study. BMC Medicine 11, 1–11 (2013).

24 Davis, K. A. et al. Indicators of mental disorders in UK Biobank—A comparison of approaches. International Journal of Methods in Psychiatric Research 28, e1796 (2019).

25 Fabbri, C. et al. Genetic and clinical characteristics of treatment-resistant depression using primary care records in two UK cohorts. Molecular Psychiatry (2021).

26 Coleman, J. R. et al. Genome-wide gene-environment analyses of major depressive disorder and reported lifetime traumatic experiences in UK Biobank. Molecular psychiatry 25, 1430–1446 (2020).

27 Manichaikul, A. et al. Robust relationship inference in genome-wide association studies. Bioinformatics 26, 2867–2873 (2010).

28 Warren, H. R. et al. Genome-wide association analysis identifies novel blood pressure loci and offers biological insights into cardiovascular risk. Nature genetics 49, 403–415 (2017).

29 Abraham, G., Qiu, Y. & Inouye, M. FlashPCA2: principal component analysis of Biobank-scale genotype datasets. Bioinformatics (2017).

30 Choi, S. W. & O’Reilly P.F. PRSice-2: Polygenic Risk Score software for biobank-scale data. GigaScience 8, doi:10.1093/gigascience/giz082 (2019).

31 Benjamini, Y. & Hochberg, Y. Controlling the false discovery rate: a practical and powerful approach to multiple testing. Journal of the Royal Statistical Society: Series B (Methodological) 57, 289–300 (1995).

32 Ridout, K. K., Ridout, S. J., Price, L. H., Sen, S. & Tyrka, A. R. Depression and telomere length: A meta-analysis. Journal of Affective Disorders 191, 237–247, doi:https://doi.org/10.1016/j.jad.2015.11.052 (2016).

33 Schutte, N. S. & Malouff, J. M. THE ASSOCIATION BETWEEN DEPRESSION AND LEUKOCYTE TELOMERE LENGTH: A META-ANALYSIS. Depression and Anxiety 32, 229–238, doi:https://doi.org/10.1002/da.22351 (2015).

34 Malouff, J. M. & Schutte, N. S. A meta-analysis of the relationship between anxiety and telomere length. Anxiety, Stress, & Coping 30, 264–272, doi:10.1080/10615806.2016.1261286 (2017).

35 Colpo, G. D. et al. Is bipolar disorder associated with accelerating aging? A meta-analysis of telomere length studies. Journal of Affective Disorders 186, 241–248, doi:https://doi.org/10.1016/j.jad.2015.06.034 (2015).

36 Huang, Y.-C., Wang, L.-J., Tseng, P.-T., Hung, C.-F. & Lin, P.-Y. Leukocyte telomere length in patients with bipolar disorder: an updated meta-analysis and subgroup analysis by mood status. Psychiatry Research 270, 41–49 (2018).

37 Joo, E.-J., Ahn, Y. M., Park, M. & Kim, S. A. Significant Shortening of Leukocyte Telomere Length in Korean Patients with Bipolar Disorder 1. Clin Psychopharmacol Neurosci 19, 559–563, doi:10.9758/cpn.2021.19.3.559 (2021).

38 Martinsson, L. et al. Long-term lithium treatment in bipolar disorder is associated with longer leukocyte telomeres. Translational Psychiatry 3, e261–e261, doi:10.1038/tp.2013.37 (2013).

39 Powell, T. R., Dima, D., Frangou, S. & Breen, G. Telomere Length and Bipolar Disorder. Neuropsychopharmacology 43, 445–453, doi:10.1038/npp.2017.125 (2018).

40 Palmos, A. B. et al. Genetic Risk for Psychiatric Disorders and Telomere Length. Frontiers in Genetics 9, doi:10.3389/fgene.2018.00468 (2018).

41 Ghimire, S., Hill, C. V., Sy, F. S. & Rodriguez, R. Decline in telomere length by age and effect modification by gender, allostatic load and comorbidities in National Health and Nutrition Examination Survey (1999-2002). PLOS ONE 14, e0221690, doi:10.1371/journal.pone.0221690 (2019).

42 Verhoeven, J. E., Penninx, B.W.J.H. & Milaneschi, Y. Unraveling the association between depression and telomere length using genomics. Psychoneuroendocrinology 102, 121–127, doi:https://doi.org/10.1016/j.psyneuen.2018.11.029 (2019).

43 Vasconcelos-Moreno, M. P. et al. Telomere Length, Oxidative Stress, Inflammation and BDNF Levels in Siblings of Patients with Bipolar Disorder: Implications for Accelerated Cellular Aging. International Journal of Neuropsychopharmacology 20, 445–454, doi:10.1093/ijnp/pyx001 (2017).

44 Gotlib, I. et al. Telomere length and cortisol reactivity in children of depressed mothers. Molecular psychiatry 20, 615 (2015).

45 Schaakxs, R., Verhoeven, J. E., Oude Voshaar, R. C., Comijs, H. C. & Penninx, B. W. J. H. Leukocyte Telomere Length and Late-Life Depression. The American Journal of Geriatric Psychiatry 23, 423–432, doi:https://doi.org/10.1016/j.jagp.2014.06.003 (2015).

46 Vance, M. C. et al. Prospective association between major depressive disorder and leukocyte telomere length over two years. Psychoneuroendocrinology 90, 157–164, doi:https://doi.org/10.1016/j.psyneuen.2018.02.015 (2018).

47 Shalev, I. et al. Internalizing disorders and leukocyte telomere erosion: a prospective study of depression, generalized anxiety disorder and post-traumatic stress disorder. Molecular Psychiatry 19, 1163–1170, doi:10.1038/mp.2013.183 (2014).

48 Chang, S.-C. et al. Prospective association of depression and phobic anxiety with changes in telomere lengths over 11 years. Depression and Anxiety 35, 431–439, doi:https://doi.org/10.1002/da.22732 (2018).

49 Wium-Andersen, M. K., Ørsted, D. D., Rode, L., Bojesen, S. E. & Nordestgaard, B. G. Telomere length and depression: Prospective cohort study and Mendelian randomisation study in 67 306 individuals. British Journal of Psychiatry 210, 31–38, doi:10.1192/bjp.bp.115.178798 (2017).

50 Demanelis, K. et al. Determinants of telomere length across human tissues. Science 369, eaaz6876, doi:doi:10.1126/science.aaz6876 (2020).

