## Supplement for "Telomere length and its associations with mental disorders, age and genetic risk for mental disorders"

This material accompanies the article

GWAS summary statistics

**Supplementary Table 1.** GWAS summary statistics

| **Phenotype** | **GWAS reference** | **Sample size** | |
|---|---|---|---|
| | | Cases | Controls |
| Anxiety disorders | Otowa et al. (2016)<br>doi: 10.1038/mp.2015.197 | 7016 | 14745 |
| Major depressive disorder | Wray et al. (2018)*<br>doi: 10.1038/s41588-018-0090-3 | 45591 | 97674 |
| Bipolar disorder | Stahl et al. (2019)<br>doi: 10.1038/s41588-019-0397-8 | 20352 | 31358 |

*Note:* GWAS = genome wide association study. *Summary statistics from Wray et al. (2018) excluding UK Biobank and 23andMe data.

T/S ratio in individuals with mental disorders

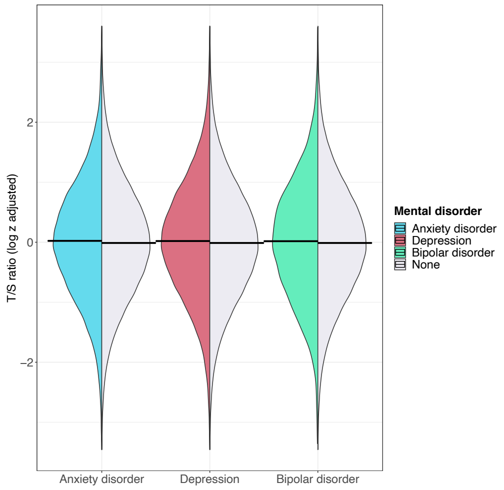

**Supplement Figure 1.** Average T/S ratio (log z adjusted) in individuals with mental disorders and non-psychiatric controls. Horizontal lines show group means. T/S ratio values below the 0.01st or above the 99.99th percentile not shown.

T/S ratio in individuals with mental disorders by antidepressant medication

Figure: T/S ratio by antidepressant medication

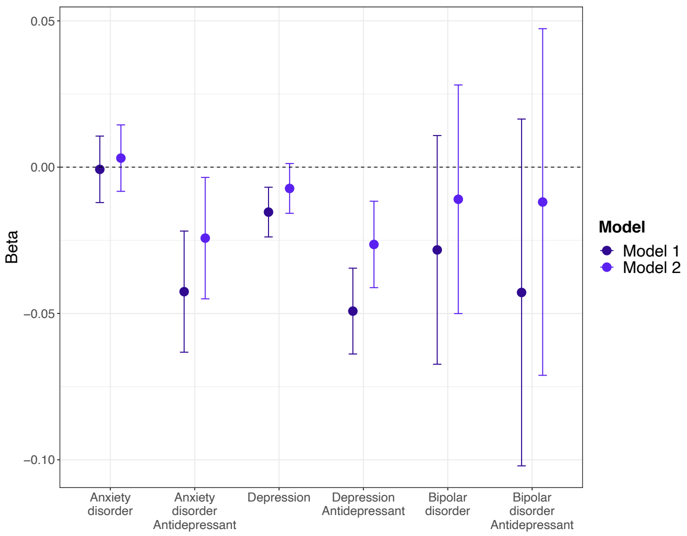

**Supplement Figure 2.** Average T/S ratio (log z adjusted) in individuals with mental disorders compared to non-psychiatric controls (reference group) stratified by antidepressant medication use. Estimates shown are ordinary least squares regression beta coefficients and 95% confidence intervals. Model 1 – adjusted for age and sex; Model 2 – adjusted for age, sex, white blood cell count, Townsend deprivation index, physical activity, smoking status, body mass index, body fat percentage and C-reactive protein.

Table: T/S ratio by antidepressant medication

**Supplement Table 2.** T/S ratio (log z adjusted) in individuals with mental disorders by antidepressant medication

| | Model 1 | | | | | Model 2 | | | | |
|---|---|---|---|---|---|---|---|---|---|---|
| Term | $\beta$ | 95% CI | | $p_{Bonf.}$ | $p_{BH}$ | $\beta$ | 95% CI | | $p_{Bonf.}$ | $p_{BH}$ |
| Controls | Ref | - | - | - | - | Ref | - | - | - | - |
| Anxiety disorder | -0.001 | -0.012 | 0.011 | >0.999 | 0.898 | 0.003 | -0.008 | 0.014 | >0.999 | 0.712 |
| + Antidepressant | -0.043 | -0.063 | -0.022 | <0.001 | <0.001 | -0.024 | -0.045 | -0.004 | 0.264 | 0.053 |
| Depression | -0.015 | -0.024 | -0.007 | 0.005 | 0.001 | -0.007 | -0.016 | 0.001 | >0.999 | 0.188 |
| + Antidepressant | -0.049 | -0.064 | -0.035 | <0.001 | <0.001 | -0.026 | -0.041 | -0.012 | 0.005 | 0.001 |
| Bipolar disorder | -0.028 | -0.067 | 0.011 | >0.999 | 0.235 | -0.011 | -0.05 | 0.028 | >0.999 | 0.712 |
| + Antidepressant | -0.043 | -0.102 | 0.016 | >0.999 | 0.235 | -0.012 | -0.071 | 0.047 | >0.999 | 0.757 |

*Note:* $\beta$ = ordinary least squares regression beta coefficient; CI = confidence interval; Ref = reference group; Bonf. = Bonferroni; BH = Benjamini & Hochberg. Model 1 – adjusted for age and sex; Model 2 – adjusted for age, sex, white blood cell count, Townsend deprivation index, physical activity, smoking status, body mass index, body fat percentage and C-reactive protein. *P*-values corrected for 12 tests.

Polygenic risk scores in individuals with mental disorders

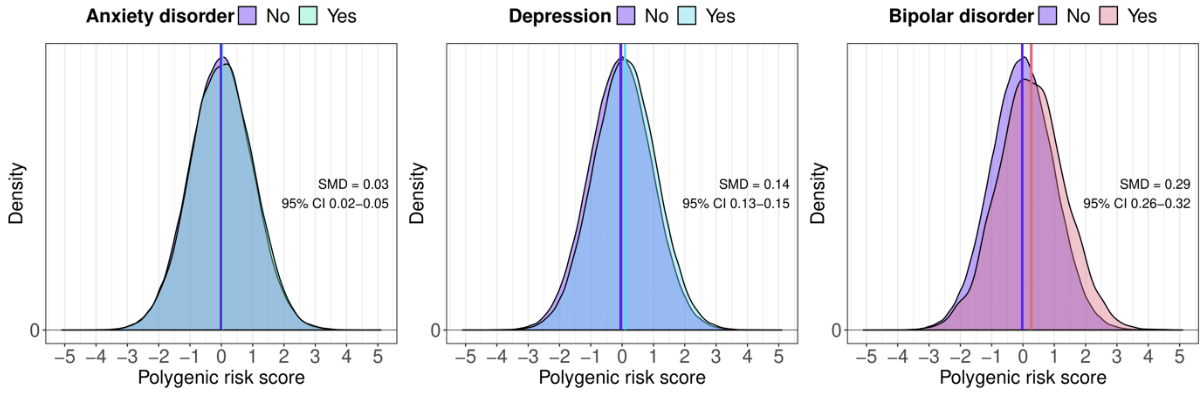

**Supplement Figure 3.** Differences in polygenic risk scores for anxiety disorder (left panel), depression (middle panel) and bipolar disorder (right panel) between individuals with these disorders and non-psychiatric controls.

Associations between T/S ratio and polygenic risk scores

Scatter plots

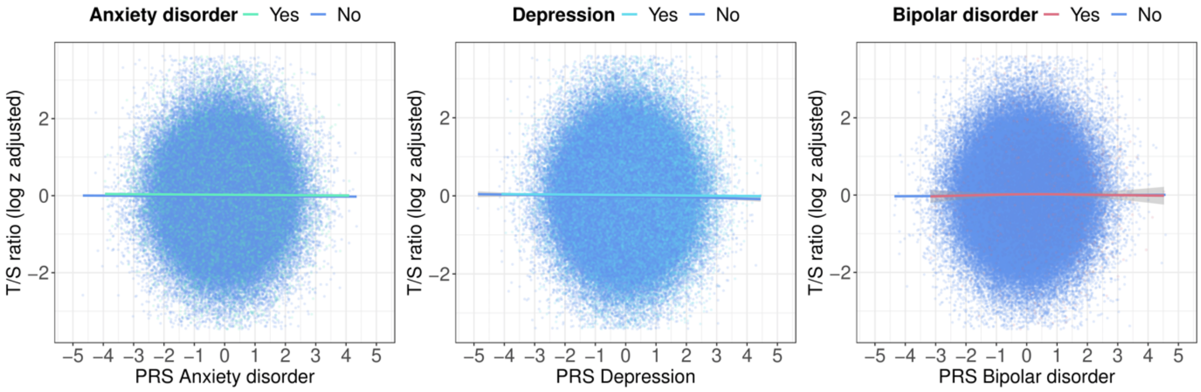

**Supplement Figure 4.** Associations between average T/S ratio (log z adjusted) and polygenic risk scores for anxiety disorder, depression and bipolar disorder. T/S ratio values below the 0.01st or above the 99.99th percentile not shown.

Table

**Supplement Table 3.** Associations between T/S ratio (log z adjusted) and polygenic risk scores for mental disorders.

| | Full sample | | | | | Cases | | | | | Controls | | | | |
|---|---|---|---|---|---|---|---|---|---|---|---|---|---|---|---|
| PRS | *β* | 95% CI | | $p_{Bonf.}$ | $p_{BH}$ | *β* | 95% CI | | $p_{Bonf.}$ | $p_{BH}$ | *β* | 95% CI | | $p_{Bonf.}$ | $p_{BH}$ |
| Anxiety disorder | -0.002 | -0.006 | 0.001 | 0.589 | 0.196 | -0.006 | -0.015 | 0.004 | >0.999 | 0.350 | -0.002 | -0.006 | 0.002 | >0.999 | 0.385 |
| Depression | -0.006 | -0.010 | -0.003 | 0.001 | 0.001 | -0.006 | -0.012 | 0.001 | 0.592 | 0.236 | -0.008 | -0.012 | -0.004 | 0.001 | 0.001 |
| Bipolar disorder | 0.003 | -0.001 | 0.008 | 0.342 | 0.171 | -0.003 | -0.036 | 0.030 | >0.999 | 0.858 | 0.003 | -0.001 | 0.008 | 0.707 | 0.236 |

*Note:* PRS = polygenic risk score; *β* = ordinary least squares regression beta coefficient; CI = confidence interval; Ref = reference group; Bonf. = Bonferroni; BH = Benjamini & Hochberg. All analyses were adjusted for the first six ancestry-informative population principal components, batch number and assessment centre. *P*-values adjusted for three (full sample) and six (cases and controls) tests.

Sensitivity analysis

Figure: T/S ratio, no comorbid depression and anxiety disorders

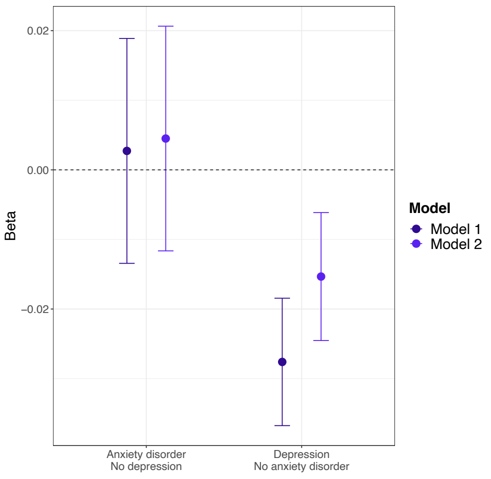

**Supplement Figure 5.** Average T/S ratio (log z adjusted) in individuals with anxiety disorders or depression compared to non-psychiatric controls (reference group), excluding individuals with comorbid anxiety disorders and depression. Estimates shown are ordinary least squares regression beta coefficients and 95% confidence intervals. Model 1 – adjusted for age and sex; Model 2 – adjusted for age, sex, white blood cell count, Townsend deprivation index, physical activity, smoking status, body mass index, body fat percentage and C-reactive protein.

Table: T/S ratio, no comorbid depression and anxiety disorders

**Supplement Table 4.** Average T/S ratio (log z adjusted), no comorbid depression and anxiety disorders.

| | Model 1 | | | | | Model 2 | | | | |
|---|---|---|---|---|---|---|---|---|---|---|
| Term | $\beta$ | 95% CI | | $p_{Bonf.}$ | $p_{BH}$ | $\beta$ | 95% CI | | $p_{Bonf.}$ | $p_{BH}$ |
| Controls | Ref | - | - | - | - | Ref | - | - | - | - |
| Anxiety disorder | 0.003 | -0.013 | 0.019 | >0.999 | 0.741 | 0.004 | -0.012 | 0.021 | >0.999 | 0.741 |
| Depression | -0.028 | -0.037 | -0.018 | <0.001 | <0.001 | -0.015 | -0.025 | -0.006 | 0.004 | 0.002 |

*Note:* $\beta$ = ordinary least squares regression beta coefficient; CI = confidence interval; Ref = reference group; Bonf. = Bonferroni; BH = Benjamini & Hochberg. Model 1 – adjusted for age and sex; Model 2 – adjusted for age, sex, white blood cell count, Townsend deprivation index, physical activity, smoking status, body mass index, body fat percentage and C-reactive protein. *P*-values corrected for four tests.

Sensitivity analysis stratified by antidepressant medication

Figure: T/S ratio, no comorbid depression and anxiety disorders by antidepressant medication

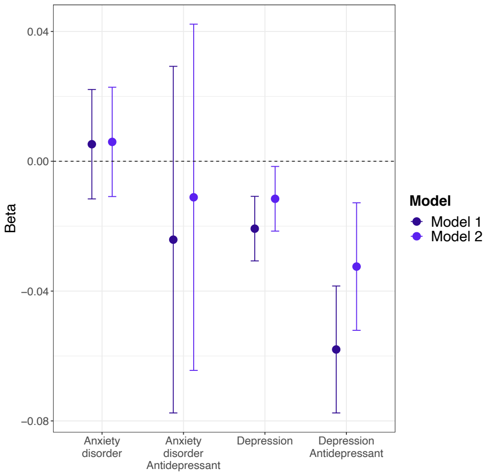

**Supplement Figure 6.** Average T/S ratio (log z adjusted) in individuals with anxiety disorders or depression compared to non-psychiatric controls (reference group) stratified by antidepressant medication use, excluding individuals with comorbid anxiety disorders and depression. Estimates shown are ordinary least squares regression beta coefficients and 95% confidence intervals. Model 1 – adjusted for age and sex; Model 2 – adjusted for age, sex, white blood cell count, Townsend deprivation index, physical activity, smoking status, body mass index, body fat percentage and C-reactive protein.

Table: T/S ratio, no comorbid depression and anxiety disorders by antidepressant medication

**Supplement Table 5.** Average T/S ratio (log z adjusted), no comorbid depression and anxiety disorders, by antidepressant medication

| | Model 1 | | | | | Model 2 | | | | |
|---|---|---|---|---|---|---|---|---|---|---|
| Term | $\beta$ | 95% CI | | $p_{Bonf.}$ | $p_{BH}$ | $\beta$ | 95% CI | | $p_{Bonf.}$ | $p_{BH}$ |
| Controls | Ref | - | - | - | - | Ref | - | - | - | - |
| Anxiety disorder | 0.005 | -0.012 | 0.022 | >0.999 | 0.618 | 0.006 | -0.011 | 0.023 | >0.999 | 0.618 |
| + Antidepressant | -0.024 | -0.078 | 0.029 | >0.999 | 0.601 | -0.011 | -0.064 | 0.042 | >0.999 | 0.683 |
| Depression | -0.021 | -0.031 | -0.011 | <0.001 | <0.001 | -0.012 | -0.022 | -0.002 | 0.184 | 0.046 |
| + Antidepressant | -0.058 | -0.078 | -0.038 | <0.001 | <0.001 | -0.032 | -0.052 | -0.013 | 0.010 | 0.003 |

*Note:* $\beta$ = ordinary least squares regression beta coefficient; CI = confidence interval; Ref = reference group; Bonf. = Bonferroni; BH = Benjamini & Hochberg. Model 1 – adjusted for age and sex; Model 2 – adjusted for age, sex, white blood cell count, Townsend deprivation index, physical activity, smoking status, body mass index, body fat percentage and C-reactive protein. *P*-values corrected for eight tests.
